# Quantifying Oxygen Demand by Patients Hospitalized with COVID-19 at a Large Safety-Net Hospital Using Multiple Methodologies

**DOI:** 10.1101/2024.03.13.24304251

**Authors:** Sky Vanderburg, Tyler Law, Priya B. Shete, Elisabeth D. Riviello, Carolyn M. Hendrickson, Gregory D. Burns, Vivek Jain, Michael S. Lipnick

**Affiliations:** Division of Pulmonary and Critical Care Medicine, San Francisco General Hospital, University of California, San Francisco; Department of Anesthesia, University of California, San Francisco; UCSF Center for Health Equity in Surgery & Anesthesia; Division of Pulmonary, Critical Care, and Sleep Medicine, Beth Israel Deaconess Medical Center, Boston, MA; Harvard Medical School; Respiratory Care Division, Department of Anesthesia and Perioperative Care, San Francisco General Hospital, University of California, San Francisco; Division of HIV, Infectious Diseases & Global Medicine, San Francisco General Hospital, University of California, San Francisco

## Abstract

**Background:** During the COVID-19 pandemic, many facilities worldwide struggled to forecast oxygen demand, which often exceeded oxygen supply to the detriment of patient care. Accurate estimates of oxygen demand by patients with COVID-19 are scarce, and proposed estimation methods have not been fully evaluated or implemented. To address this knowledge gap, oxygen demand by COVID-19 patients was calculated at a large safety-net hospital in the United States using patient consumption (demand) data, oxygen procurement (supply) data, and modeled data with a novel calculator tool.

**Methods:** Data were extracted from electronic medical records of patients admitted with COVID-19 to Zuckerberg San Francisco General Hospital (ZSFG) from March 2020 to March 2022, including every recorded peripheral oxygen saturation (SpO_2_) measurement as well as oxygen delivery device(s) and settings. Total patient oxygen consumption was calculated as the sum of oxygen delivery amounts for each recorded time interval during hospitalization. Oxygen delivery amounts were calculated using delivery device-specific formulas. Patient and treatment-specific factors which may impact oxygen demand were also reported. For comparison, oxygen procurement logs from the study period were reviewed to estimate supply consumed, and the Oxygencalculator.com tool was used to model oxygen demand using an experimental patient population of the same size.

**Results:** In total, 282,095 time points from 1,076 patients were analyzed. Two-thirds of patients received oxygen, of which 24.3% received high-flow oxygen (HFO) therapy and 16.0% received invasive mechanical ventilation (IMV) at some point. In-hospital mortality was 7.5% overall, 10.8% for patients who received oxygen, and 28.3% for patients who received IMV. The median (IQR) duration of oxygen therapy was 3.1 (0.8-8.9) days, mean (SD) oxygen flow was 5.6 (5.0) liters per minute (LPM), and mean (SD) total volume of oxygen delivered was 180,115 (510,330) liters (L) per hospitalization. Both the supply- and model-based methods overestimated oxygen consumption compared to demand estimated from patient data.

**Conclusions:** This study represents one of the largest cohorts of patients with COVID-19 for which oxygen demand has been calculated, including patient clinical characteristics which may help explain variations in oxygen demand. Moreover, oxygen demand was quantified using a methodology that could be applied in any setting.

## BACKGROUND

Oxygen therapy is an essential treatment for hypoxemia due to COVID-19 and other causes of acute hypoxemic respiratory failure (AHRF). Despite oxygen’s listing as an Essential Medicine by the World Health Organization (WHO), longstanding oxygen supply limitations have plagued healthcare facilities in many low and middle-income countries (LMICs).^1-8^ During the COVID-19 pandemic, periodic and regional increases in demand for oxygen strained or exceeded supply in many more locations worldwide, including hospitals in high-income countries.^9-11^

Throughout the pandemic, decision makers at every level—clinicians, facilities, ministries of health, and aid organizations—struggled to forecast oxygen needs. This was due in no small part to lack of data on oxygen requirements by patients with COVID-19, an uncertainty which was compounded by factors such as varying infectivity and pathogenicity of COVID-19 strains as well as regional differences in respiratory support equipment and practices. These knowledge gaps— often combined with oxygen supply limitations—resulted in oxygen shortages which negatively impacted patient care.

Of the many reports of oxygen supply shortages and/or strains during the COVID-19 pandemic, few attempted to quantify oxygen demand.^12,13^ Importantly, none quantify oxygen demand at the facility level or over the course of multiple surges of COVID-19 cases. Estimates of facility-level oxygen demand are crucial to optimizing oxygen delivery capacity for three reasons. First, they improve upon non-empirical estimations of oxygen demand estimates currently used by most local, national, and international stakeholders to guide planning.^14-16^ Second, facility-level estimations can explicitly incorporate factors that vary across facilities and over time, such as COVID-19 strains, types of oxygen delivery devices used, and prevailing oxygen titration practices. Third, these estimates provide a quantitative benchmark to assess the impact of facility-level interventions such as quality improvement initiatives to conserve oxygen.

Without more accurate estimates of facility-level oxygen demand and contributing factors, our understanding of oxygen delivery for COVID-19 and our preparedness for the next pandemic will be incomplete. The aim of this study is threefold: 1) to describe key demographic and clinical characteristics of a large cohort of patients hospitalized with COVID-19 at a safety-net hospital, 2) to estimate oxygen demand in this cohort using patient-level oxygen delivery data; and 3) to compare oxygen demand estimates with those derived from oxygen procurement data and an open-source tool for modeling oxygen demand.

## METHODS

### Study Population and Setting

For this retrospective cohort study, data was extracted from the electronic medical records (EMR) of all patients admitted with COVID-19 at Zuckerberg San Francisco General Hospital (ZSFG) from March 1, 2020 through March 1, 2022. COVID-19 illness was defined by the identification of SARS-CoV-2 virus by polymerase chain reaction (PCR) in nasopharyngeal samples obtained during the coinciding hospitalization. ZSFG is a large public hospital in San Francisco, California, with 284 total licensed beds and 16 operating theatres. Of the total beds, 58 and 13 are in adult and neonatal tensive care units (ICUs), respectively, while 184 are in adult, 8 in pediatric, and 21 are in perinatal wards, respectively. Pediatric trauma patients are cared for in the ZSFG ICU, though medical ICU patients are transferred to a dedicated pediatric hospital. ZSFG is the primary provider of safety-net health care for San Francisco, providing care for >100,000 patients annually, 80% of which receive publicly funded health insurance or are uninsured.

Hypoxemic patients with COVID-19 received standard oxygen delivery modalities at ZSFG, consisting of standard flow oxygen (SFO), high flow oxygen (HFO), and mechanical ventilation for hypoxemic patients. Peripheral oxygen saturation (SpO_2_) monitoring was done using Signal Extraction Technology^®^ (SET^®^) finger and earlobe probes (Masimo Corporation, Irvine, CA) and IntelliVue patient monitors (Philips Healthcare, Andover, MA). HFO was used in both the medical wards and ICU. Patients with COVID-19 requiring ICU-level care were co-located in two 10-bed “COVID-19 ICUs” staffed by dedicated physician, nurse, and respiratory therapist teams. Though the treatment of COVID-19 evolved to include anti-viral, corticosteroid, and other immunomodulatory therapies during the study period, there were no explicit departures—such as policies requiring early intubation—from pre-existing standards of care for AHRF and acute respiratory distress syndrome (ARDS). The primary source of oxygen at ZSFG is liquid oxygen (LOX) which is then piped in gaseous form to the patient’s room. No shortages or rationing of oxygen supply or other respiratory support resources (e.g., mechanical ventilators) occurred at this hospital during the COVID-19 pandemic.

### Data Extraction

Data were extracted through a computerized search of each patient’s EMR, including vital signs, select laboratory tests, elements of the Universal Vital Assessment (UVA) score, respiratory support, COVID-19 treatments, and outcomes. The resulting dataset represented all time points where ≥1 variable of interest was recorded through automated or manual capture over the course of the patient’s entire hospitalization, including every recorded SpO_2_ measurement, oxygen therapy titration, and oxygen delivery device. Data extraction was done in consultation with the Clinical and Translational Science Institute at University of California San Francisco (UCSF) and was de-identified before provision to study investigators for analysis. This study protocol was reviewed and granted approval and waiver of individual patient consent by the UCSF Institutional Review Board and ZSFG Administration.

### Definitions

COVID-19 severity was classified as follows: 1) “moderate” for hospitalized patients who did not receive supplemental oxygen; 2) “severe” for hospitalized patients who received supplemental oxygen and/or non-invasive ventilation (NIV) but not IMV; and 3) “critical” for hospitalized patients who received IMV. These categories align with classifications of COVID-19 severity recommended by the WHO.^15^ The universal vital assessment (UVA) score was also used to describe severity of critical illness and facilitate comparisons with cohort worldwide, as the UVA uses variables that are routinely collected worldwide as part of clinical care. UVA score >4 indicates higher odds of death compared to UVA score ≤1 (odds ratio (OR) 10.41).^17^

The earliest recorded values of variables were used to classify COVID-19 and hypoxemia severity and assign UVA scores. When partial pressure of oxygen (PaO2) values were not available, Non-linear imputation of arterial PaO_2_ from SpO_2_ was used to calculate imputed PaO_2_/FiO_2_ ratios for cases where SpO_2_ <97%; imputation of PaO_2_ from SpO_2_ ≥97% is unreliable.^18^ At time points where FiO_2_ was not recorded for SFO (i.e., nasal cannula, facemask, or facemask with nonrebreather), it was estimated from oxygen flow (LPM) using the following previously described formula: FiO_2_=0.21+0.03(O_2_ LPM).^19,20^ Otherwise, time points missing FiO_2_ recordings were excluded. Severe hypoxemia was defined by PaO_2_/FiO_2_ ratio <150, SpO_2_/FiO_2_ ratio <270, or imputed PaO_2_/FiO_2_ ratio <150.^18,21,22^

### Analysis

Oxygen delivery flows (liters per minute) for each time interval were first calculated using formulas derived for each type of oxygen delivery device in use (Table S1). Oxygen delivery flows were assumed to be constant for the time interval between consecutive oxygen delivery recordings. Oxygen delivery flows for each time interval were then multiplied by the length of the time interval (milliseconds) to obtain oxygen delivery amounts (L). Missing values for variables required to calculate oxygen delivery flows were imputed using the immediately preceding recorded value in the time series, since most time intervals were short (milliseconds to minutes). Total oxygen demand for each patient’s hospitalization was calculated using the following previously described formula: ∑_*n*_ *X*_*n*_ (*T*_(*n*+1)_ − *T*_*n*_), where *X*_*n*_= oxygen delivery flow and (*T*_(*n*+1)_ − *T*_*n*_)= the time interval between oxygen delivery recordings (milliseconds).

The mean oxygen flow in LPM for each patient receiving oxygen therapy was calculated as their total oxygen demand divided by the duration of oxygen therapy. Additional sub-analyses of total volume (L) and mean oxygen flow in LPM delivered via each oxygen delivery device were also conducted. Median and interquartile range (IQR) values were reported where data were non-normally distributed. Analyses were conducted using STATA/SE 18.0 (StataCorp LLC).

### Comparison with Other Estimation Methods

Procurement data for the liquid oxygen (LOX) tank at ZSFG during the study period was used to approximate the monthly total volume of gaseous oxygen procured for the entire facility using standard conversion factors—which was then divided by 30 to estimate the daily total demand. For the model-based estimation method, the open access oxygencalculator.com tool was used which models the oxygen consumption of a facility with a given number of patients under five different COVID-19 severity scenarios.^23^ The proportion of patients using different types of oxygen support (Table S2) is varied according to the severity assumptions (e.g., 4% using HFO in Scenario 1 versus 26% in Scenario 5). Daily oxygen consumption under Scenario 3—the “default” scenario—was estimated using the same number of patients and mean oxygen delivery device settings derived from EMR data. Where possible, oxygen demand estimates by different estimation methods were compared using Student’s t test.

## RESULTS

In total, 282,095 time points were analyzed from 1,076 patients whose demographic and clinical characteristics are summarized in Table 1. The median (IQR) age (years) was 54 (37-67), and the cohort was predominantly male (60.2%), self-reported race “Other” (50.5%), and self-reported ethnicity Hispanic and/or Latino/a/x (50.5%). About one-third of patients had a body mass index (BMI) ≥30. Nearly two-thirds (63.8%) of patients met criteria for severe or critical COVID-19 disease severity during their hospitalization, and UVA score was >4 for 82 (7.6%) patients. Around one-fourth (27.0%) of patients were initially severely hypoxemic.

**Table 1.** Patient and COVID-19 Treatment Characteristics.

|  |  |
| --- | --- |
|  | N=1,076* |
| **Age (years), median (IQR)** | 54 (37-67) |
| **Male sex, n (%)** | 648 (60) |
| **Race (self-reported), n (%)** |  |
| Other<sup>†</sup> | 539 (50) |
| Black or African American | 171 (16) |
| Asian | 158 (15) |
| White | 144 (13) |
| Hawaiian or Other Pacific Islander | 37 (3) |
| Decline to Answer | 18 (2) |
| American Indian or Alaska Native | 9 (1) |
| **Ethnicity (self-reported), n (%)** |  |
| Hispanic, Latino/a/x, or Spanish origin | 543 (51) |
| **BMI, median (IQR)** | 27.6 (24-33) |
| **BMI $\geq 30$, n/N (%)** | 356/984 (36) |
| **UVA Score, median (IQR)** | 0 (0-1) |
| **UVA Score $>4$, n (%)** | 82 (8) |
| **COVID-19 Severity, n (%)** |  |
| Moderate | 341 (32) |
| Severe | 513 (48) |
| Critical | 173 (16) |
| Unknown<sup>‡</sup> | 49 (5) |
| **Initial<sup>§</sup> Hypoxemia Severity** |  |
| PaO<sub>2</sub>/FiO<sub>2</sub> Ratio, median (IQR) | 219 (143-329) |
| PaO<sub>2</sub>/FiO<sub>2</sub> Ratio <150, n/N (%) | 105/365 (29) |
| Imputed PaO<sub>2</sub>/FiO<sub>2</sub> Ratio, median (IQR) | 262 (158-337) |
| Imputed PaO<sub>2</sub>/FiO<sub>2</sub> Ratio <150, n/N (%) | 47/201 (23) |
| **Received remdesivir, n/N (%)** | 594/886 (67) |
| **Received corticosteroids, n/N (%)** | 218/886 (25) |
| **Received non-steroid immunomodulatory therapy\*\***, n/N (%) | 72/886 (8) |
| **LOS (days), median (IQR)** | 5.0 (2-10) |
| **In-hospital mortality, n/N (%)** |  |
| Overall | 81/1,076 (8) |
| Moderate COVID-19 | 7/341 (2) |
| Severe COVID-19 | 25/513 (5) |
| Critical COVID-19 | 49/173 (28) |

\*N=1,076 for all reported percentages unless otherwise specified.
†Self-reported race reported as “other” without further detail.
‡Oxygen provision and/or delivery device not recorded.
§First recorded value(s) for a given patient.
\*\*Immunomodulatory therapy=baricitinib, tofacitinib, tocilizumab, or sarilumab.

During the study period, the average daily total inpatient census was 221, of which 1-69 (<1%-31%) were patients with COVID-19. The median (IQR) length of stay (LOS) was 5.0 (2.0-10.0) days. In-hospital mortality was 7.5% for all COVID-19 patients and 2.1%, 4.9%, and 28.3% for patients with moderate, severe, and critical COVID-19, respectively. Both remdesivir and corticosteroids were given to 206 (19.1%) patients, and 50 (4.6%) patients received remdesivir, corticosteroids, and ≥1 non-steroid immunomodulatory therapy.

Characteristics of oxygen therapy delivery are summarized in Table 2. In total, 686 (63.8%) patients received oxygen at any point during their hospitalization, and the median (IQR) duration of oxygen therapy was 3.1 (0.8-8.9) days. Of these, 261 (24.3%) received HFO therapy, of which 144 (55.2%) did not require IMV. Overall, 173 (16.0%) of patients required IMV. For both groups of patients who did and did not receive oxygen therapy, the median average SpO_2_ over the course of hospitalization was the same (96.2%).

**Table 2.**
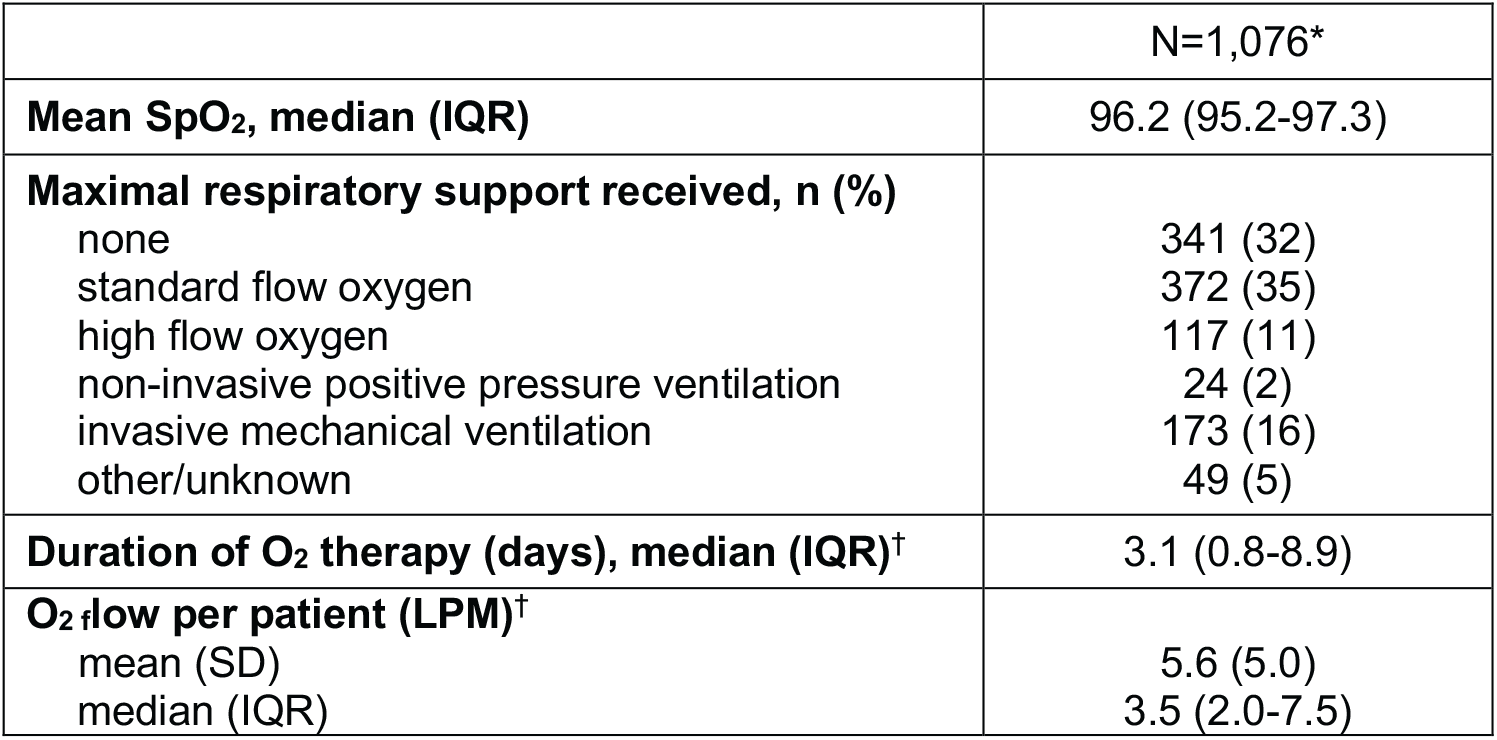
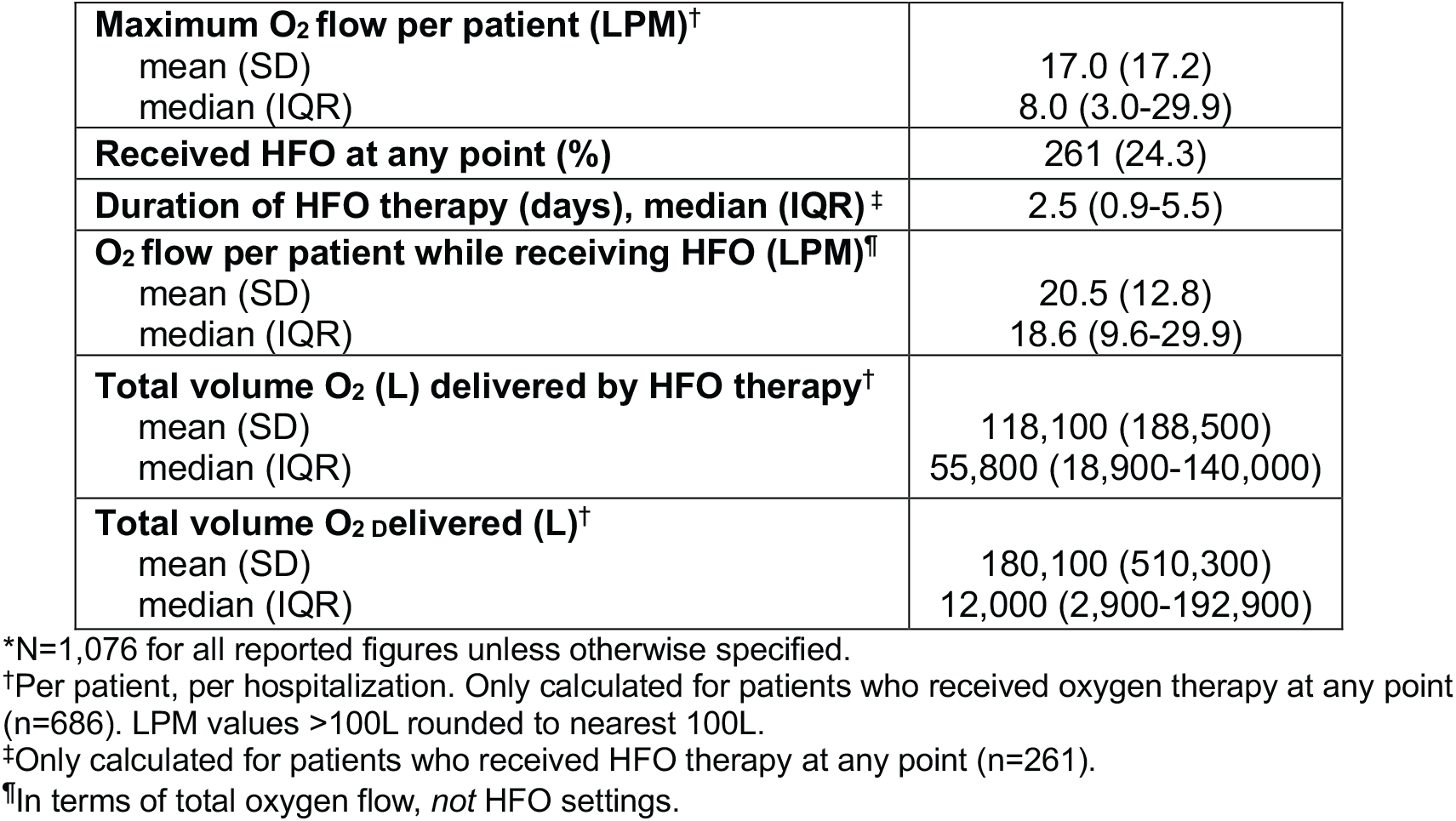
Oxygen Delivery Characteristics.

Median (IQR) maximum oxygen flow was 8.0 (3.0-29.9) LPM, and median (IQR) total volume of oxygen delivered per patient was 11,959 L (2,852-192,864) over the entire hospitalization. Median (IQR) total volume of oxygen delivered per day across all patients was 54,002 L (20,457-105,068), and the total range was 0.0-382,504 L. Daily volume of oxygen delivered and the relative contributions of each type of oxygen delivery device over the study period is shown in Figure 1.

**Figure 1.**
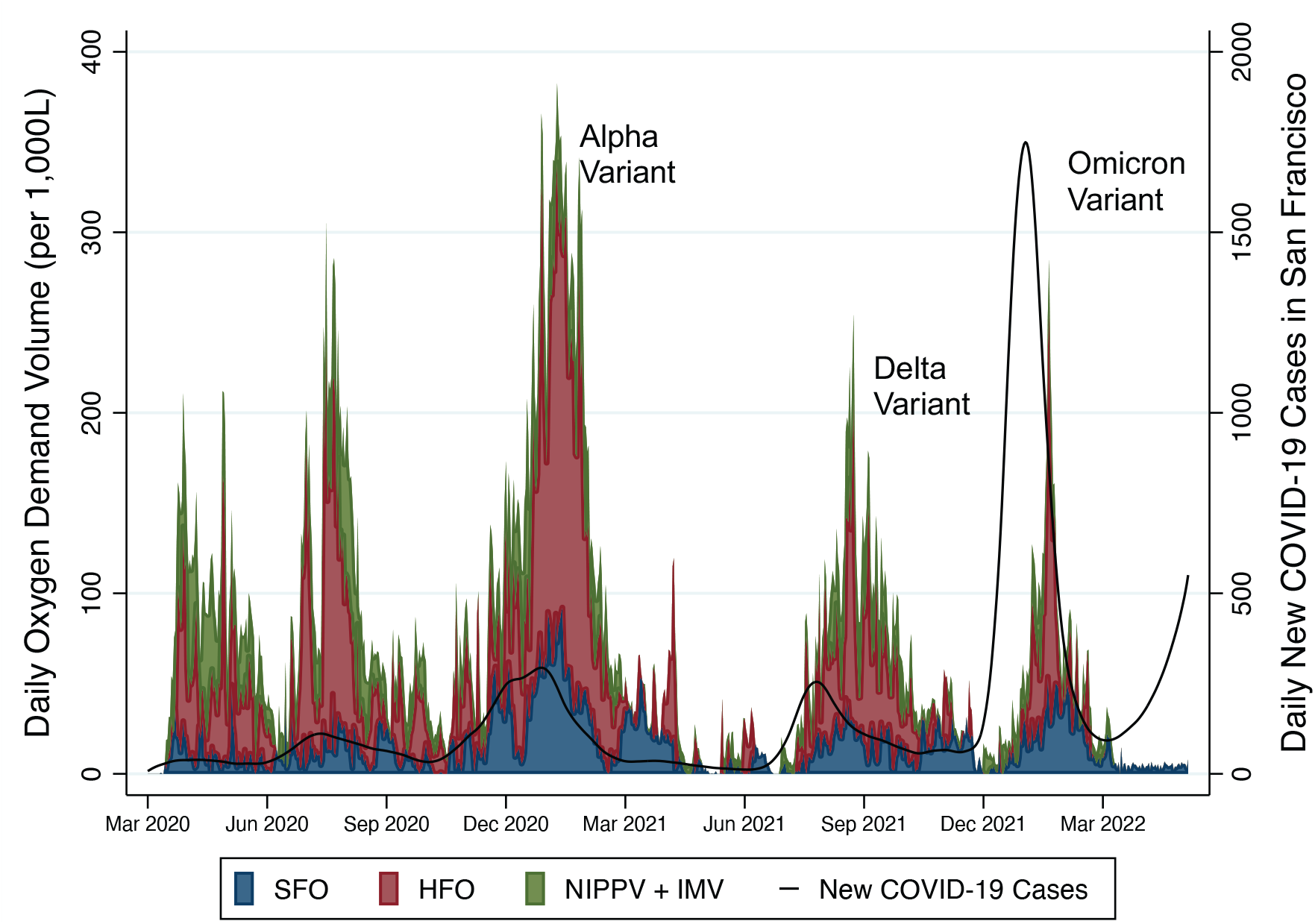
Oxygen Demand of COVID-19 Patients at Zuckerberg San Francisco General Hospital. Source of COVID-19 Case Data: Government of San Francisco (https://data.sfgov.org/; accessed June 19, 2023). *NIPPV*, non-invasive positive pressure ventilation

Overall, HFO therapy accounted for the largest component of oxygen delivered. On average, HFO therapy accounted for 47.9% of the total daily volume of oxygen delivered on days when HFO therapy was being delivered to patients with COVID-19 (i.e., excluding days when no oxygen was delivered by HFO therapy). The mean (SD) flow and FiO_2_ setting for HFO therapy was 38.9 LPM (6.7) and 0.61 (0.23), respectively. On average, SFO therapy accounted for 37.5% of the total daily volume of oxygen delivered on days when SFO therapy was delivered.

Estimates of oxygen demand based on LOX procurement and calculator-based modeling are presented in Figures 2 and 3, respectively. Compared to LOX delivery during the immediately preceding 30-month period, the mean total daily volume of oxygen delivered during the study period was significantly higher (p<0.01; 1.594 to 10.615 million liters more per month). Calculator-based modeling also yielded higher estimates in volume of oxygen demand, with a mean daily difference of 93,583 liters (74.186 million liters more over study period).

**Figure 2.**
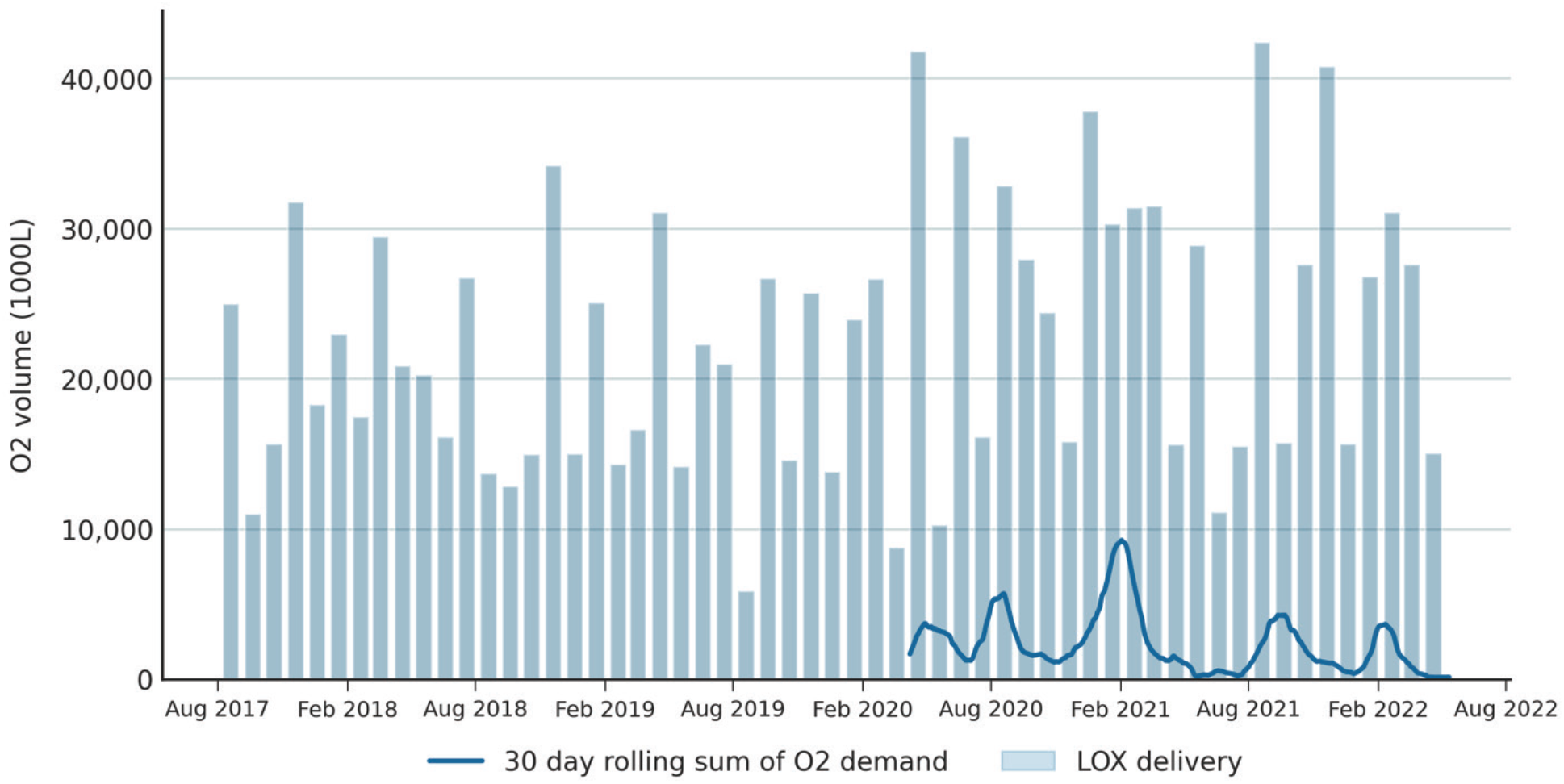
Estimation Using Patient-level Oxygen Consumption versus LOX Procurement Data.

**Figure 3.**
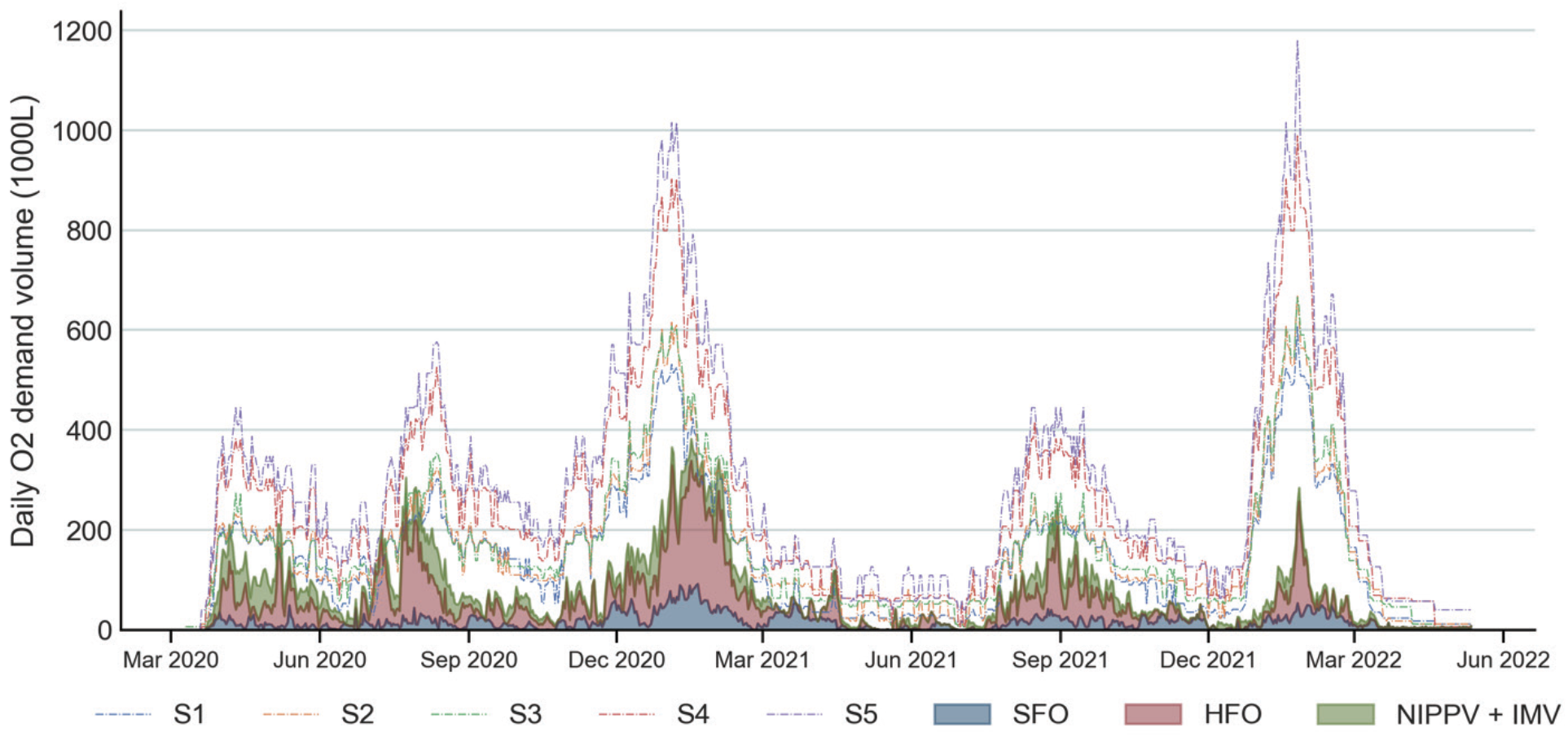
Estimation Using Patient-level Oxygen Consumption versus Model Scenarios. S1-S5 represent severity scenarios used by the oxygencalculator.com tool to vary the proportion of oxygen delivery devices used in a given population of patients, with S1 and S5 representing the least and most severe, respectively.

## DISCUSSION

Here for the first time we compare quantification of oxygen demand using various methodologies applied to a large retrospective study of patients hospitalized with COVID-19 during the first two years of the pandemic. These estimates provide critical insights into the true oxygen demand by patients hospitalized with COVID-19 treated in accordance with National Institutes of Health (NIH) guidelines^24^ in a setting that was not impacted by rationing of care or resources. Moreover, our study is one of the largest cohorts of patients with COVID-19 for which oxygen demand estimates have been derived and one of the first to compare various methodologies for quantifying oxygen usage.

Subjects in this cohort were predominately young, male, and reported race as non-white. Roughly half reported Hispanic ethnicity. Most patients (67%) received remdesivir, but only one quarter of patients received corticosteroid therapy. It should be noted that many patients were enrolled before the publication of the RECOVERY trial which changed practice to incorporate standard use of corticosteroids in patients hospitalized with COVID-19 and requiring supplemental oxygen.^25^ This may reflect patient data prior to widespread use of corticosteroid therapy for COVID-19, or it may be due to variations in practices of recording COVID-19 therapies in the EMR. For the 63.8% of patients who received supplemental oxygen (i.e., severe or critical COVID-19), the mean flow, median total duration, and mean total volume of oxygen delivered to each patient was 5.6 LPM, 3.1 days, and 180,115 L, respectively.

Although the peaks in oxygen demand estimated by the supply- and model-based approaches roughly correspond to peaks in oxygen demand estimated by the EMR-based approach, they largely overestimated oxygen demand by comparison. Much of the difference between supply- and EMR-based approaches can be attributed to other sources of oxygen consumption, namely oxygen provision for patients without COVID-19 and leak/wastage. The daily equivalent of LOX procured during the study period was 35% higher than preceding 30-month period. Differences between the model-based and EMR-based estimates likely arise from assumptions for the Oxygencalculator.com tool model, which were based on a scoping review of available data from the early case series of COVID-19 patients from China, Italy, and New York City. The largest source of variation was HFO consumption estimates, as the oxygencalculator.com tool defaults to 100% FiO_2_ and 40 LPM flow for all patients receiving HFO. Readjusting the model to reflect mean FiO_2_ and flow values for HFO found in the study cohort, more closely predicted patient demand. These data are being incorporated into the next version of the oxygencalculator.com tool. Less severe scenarios came closer to the observed demand than more severe scenarios. This was particularly pronounced during the Omicron wave, where the calculated oxygen consumption gave a significant overestimate.

Oxygen demand estimation using patient-level EMR data is relatively straightforward and dependent only on having data on oxygen delivery devices and their settings. It is a methodology that can be used for many critical roles, including the following: 1) improving upon non-empirical methods to plan oxygen supply, both in settings that need to prepare for acute potential oxygen scarcity—such as hospitals in the United States (US) during a pandemic—as well as settings with chronic oxygen scarcity, such as those in many LMICs; 2) facilitating comparisons across facilities by explicitly characterizing factors which can impact oxygen demand; 3) setting and assessing quantitative benchmarks for facility-level interventions to improve oxygen delivery. Importantly, oxygen demand estimation can be used for all oxygen therapy scenarios, not just treatment of patients with hypoxemia due to COVID-19.

Delivery amounts estimated by this study are higher than those from a Swedish study of COVID-19 patients—which is, to the best of our knowledge, the only other published estimate of oxygen delivery for COVID-19 from patient-level data.^12^ However, the Swedish cohort excluded patients with critical disease, and HFO therapy was not used. Additionally, the mean oxygen flow for our study is lower than the 10 LPM and 30 LPM estimated by WHO expert consensus for severe and critical COVID-19, respectively.^14^

There are several important factors to consider when interpreting patient-level estimates of oxygen demand. First, the study period spanned the introduction of remdesivir (April 2020), corticosteroids (June 2020), and vaccines (January 2021) as well as other anti-viral and immunomodulatory therapies, all offering proven or plausible reductions in oxygen requirements by COVID-19 patients. Second, HFO therapy—which accounted for roughly half the daily oxygen demand on average—utilizes much more oxygen than other oxygen delivery modalities when used at maximum or near-maximum device capacity.^26^ However, the volume of oxygen required for HFO therapy is highly dependent on flow and FiO_2_ settings, for which titration practices vary widely across providers and facilities.^27,28^ Third, oxygen demand per COVID-19 hospitalization decreased with the wave of cases due to the Omicron variant of the pandemic (Figure 1), correlating with decreased severity of disease which was widely reported. Fourth, the overall in-hospital mortality (7.5%) for all COVID-19 patients was lower than those reported in national and international cohorts of hospitalized COVID-19 patients from similar time periods.^29,30^ However, comparisons of mortality should be interpreted cautiously, as the distribution of patient risk factors, available resources, and practices of care varied significantly between hospitals and time periods during the pandemic. Importantly, the descriptions of treatments and patient outcomes provided by this study do not reflect any limitations or rationing of supplies undertaken by many US hospitals during regional surges in COVID-19 hospitalizations. Finally, the quantification methodology for oxygen demand described herein used EMR data with frequent timepoints which may not be available in all healthcare settings. However, the same validated equations for oxygen utilization could be leveraged with manually recorded oxygen delivery device data, as is currently being done in a study of hypoxemic patients at five hospitals in sub-Saharan Africa (ClinicalTrials.gov NCT05754034; https://clinicaltrials.gov/study/NCT05754034).

Our study has several limitations. First, these estimates are from a single center. We were, however, able to describe many patient characteristics that impact oxygen demand, providing descriptive context for the oxygen quantities used. Second, given the retrospective nature of the study, we can only adjudicate the amount of oxygen patients received, not the amount they required; however, there was no oxygen resource scarcity and the mean SpO_2_ was within the recommended range. Similarly, we cannot verify that oxygen delivery amounts did not change between recorded time intervals; however, time intervals were very short (on average, 64.7 minutes). Lastly, these are estimates in COVID-19 patients during a time of wide variability in disease severity due to changes in practice and pathogen virulence. Generalization of these findings to patients with all causes of hypoxemia should be done with caution.

## CONCLUSIONS

This study represents one of the largest cohorts of patients with COVID-19 for which oxygen demand has been calculated. Because this study was done in a setting that did not experience rationing of care or deviations from pre-pandemic respiratory care protocols, it may be the best available estimate of oxygen demand by COVID-19 patients. This primary methodology for demand estimation could be relatively easily reproduced in settings with an EMR, whereas the modeling tool, if refined and validated further, could provide a rapid method for use in all settings to help plan for acute or chronic crises in oxygen demand.

## Data Availability

The datasets used and/or analyzed during the current study are available from the corresponding authors on reasonable request.

https://www.oxygencalculator.com/oxygen/o2demand

## Acknowledgements

Data extraction was performed by staff from the Clinical and Translational Science Institute at UCSF and funded by the UCSF Center for Health Equity in Surgery and Anesthesia. Guidance for data extraction was given by Jen Berke, Informatics Liaison for San Francisco General Hospital. The oxygen calculator (oxygencalculator.com) was developed with funds from the USAID-STAR project.

## LIST OF ABBREVIATIONS

COVID-19: 
LMIC: 
ZSFG: 
SARS-CoV-2: 
LPM: 
SFO: 
HFO: 
NIPPV: 
IMV: 
PaO_2_: 
SpO_2_: 
FiO_2_: 
UVA: 
BMI: 
LOS: 
IQR: 
OR: 
SD: 
WHO: 
NIH: 
FDA: 

## DECLARATIONS

This study protocol was reviewed and granted approval and waiver of individual patient consent by the UCSF Institutional Review Board and ZSFG Administration.

### Availability of data and materials

The datasets used and/or analyzed during the current study are available from the corresponding author on reasonable request.

## REFERENCES

1. World Health Organization. WHO model list of essential medicines - 22nd list. 2021. 30 September 2021. https://www.who.int/publications/i/item/WHO-MHP-HPS-EML-2021.02

2. World Health Organization Executiive Board. Increasing access to medical oxygen: Draft decision proposed by Australia, Bangladesh, Central African Republic, European Union and its 27 Member States, Kenya, Türkiye and Uganda. 2023;

3. Meara JG, Leather AJ, Hagander L, et al. Global Surgery 2030: evidence and solutions for achieving health, welfare, and economic development. Lancet. Aug 8 2015;386(9993):569–624. doi:10.1016/S0140-6736(15)60160-X

4. Sutherland T, Musafiri S, Twagirumugabe T, Talmor D, Riviello ED. Oxygen as an Essential Medicine: Under- and Over-Treatment of Hypoxemia in Low- and High-Income Nations. Crit Care Med. Oct 2016;44(10):e1015–6. doi:10.1097/CCM.0000000000001912

5. Belle J, Cohen H, Shindo N, et al. Influenza preparedness in low-resource settings: a look at oxygen delivery in 12 African countries. J Infect Dev Ctries. Aug 4 2010;4(7):419–24. doi:10.3855/jidc.859

6. Graham HR, Olojede OE, Bakare AA, et al. Measuring oxygen access: lessons from health facility assessments in Lagos, Nigeria. BMJ Glob Health. Aug 2021;6(8)doi:10.1136/bmjgh-2021-006069

7. Mangipudi S, Leather A, Seedat A, Davies J. Oxygen availability in sub-Saharan African countries: a call for data to inform service delivery. Lancet Glob Health. Sep 2020;8(9):e1123–e1124. doi:10.1016/S2214-109X(20)30298-9

8. African C-CCOSI. Patient care and clinical outcomes for patients with COVID-19 infection admitted to African high-care or intensive care units (ACCCOS): a multicentre, prospective, observational cohort study. Lancet. May 22 2021;397(10288):1885–1894. doi:10.1016/S0140-6736(21)00441-4

9. Craxi L, Vergano M, Savulescu J, Wilkinson D. Rationing in a Pandemic: Lessons from Italy. Asian Bioeth Rev. Sep 2020;12(3):325–330. doi:10.1007/s41649-020-00127-1

10. Lin R-G, Karlamangla S, Money L. L.A. County hospitals running dangerously low on oxygen, supplies as ER units are overwhelmed. Los Angeles Tiimes. 2020. Accessed 25 December 2020.

11. Holmes K, Elamroussi A. First, surges in Covid-19 infections led to shortages of hospital beds and staff. Now it’s oxygen. CNN; 2021. 30 August 2021. https://www.cnn.com/2021/08/29/health/us-coronavirus-sunday/index.html?utm_source=newsletter&utm_medium=email&utm_campaign=newsletter_axiosvitals&stream=top

12. Hvarfner A, Al-Djaber A, Ekstrom H, et al. Oxygen provision to severely ill COVID-19 patients at the peak of the 2020 pandemic in a Swedish district hospital. PLoS One. 2022;17(1):e0249984. doi:10.1371/journal.pone.0249984

13. White HD, Danesh V, Ogola GO, Jimenez EJ, Arroliga AC. Quantifying oxygen supply and demand during the COVID-19 pandemic: An integrated health system perspective. Intensive Crit Care Nurs. Apr 2023;75:103374. doi:10.1016/j.iccn.2022.103374

14. World Health Organization. Oxygen sources and distribution for COVID-19 treatment centres: Interim guidance. 2020. 4 April 2020. https://www.who.int/publications/i/item/oxygen-sources-and-distribution-for-covid-19-treatment-centres

15. World Health Organization. Living Guidance for Clinical Management of COVID-19. 2021. 23 November 2021. https://www.who.int/publications/i/item/WHO-2019-nCoV-clinical-2021-2

16. World Health Organization. COVID-19 Essential Supplies Forecasting Tool, version 4.1. 2022. Accessed 15 February 2022. https://www.who.int/publications/i/item/WHO-2019-nCoV-Tools-Essential_forecasting-2022.1

17. Moore CC, Hazard R, Saulters KJ, et al. Derivation and validation of a universal vital assessment (UVA) score: a tool for predicting mortality in adult hospitalised patients in sub-Saharan Africa. BMJ Glob Health. 2017;2(2):e000344. doi:10.1136/bmjgh-2017-000344

18. Brown SM, Duggal A, Hou PC, et al. Nonlinear Imputation of PaO2/FIO2 From SpO2/FIO2 Among Mechanically Ventilated Patients in the ICU: A Prospective, Observational Study. Crit Care Med. Aug 2017;45(8):1317–1324. doi:10.1097/CCM.0000000000002514

19. Coudroy R, Frat JP, Girault C, Thille AW. Reliability of methods to estimate the fraction of inspired oxygen in patients with acute respiratory failure breathing through non-rebreather reservoir bag oxygen mask. Thorax. Sep 2020;75(9):805–807. doi:10.1136/thoraxjnl-2020-214863

20. Frat JP, Quenot JP, Badie J, et al. Effect of High-Flow Nasal Cannula Oxygen vs Standard Oxygen Therapy on Mortality in Patients With Respiratory Failure Due to COVID-19: The SOHO-COVID Randomized Clinical Trial. JAMA. Sep 27 2022;328(12):1212–1222. doi:10.1001/jama.2022.15613

21. Guerin C, Reignier J, Richard JC, et al. Prone positioning in severe acute respiratory distress syndrome. N Engl J Med. Jun 6 2013;368(23):2159–68. doi:10.1056/NEJMoa1214103

22. Chen W, Janz DR, Shaver CM, Bernard GR, Bastarache JA, Ware LB. Clinical Characteristics and Outcomes Are Similar in ARDS Diagnosed by Oxygen Saturation/Fio2 Ratio Compared With Pao2/Fio2 Ratio. Chest. Dec 2015;148(6):1477–1483. doi:10.1378/chest.15-0169

23. Oxygen Calculator. oxygencalculator.com. San Francisco, CA.

24. National Institutes of Health. COVID-19 Treatment Guidelines. Updated November 2, 2023. Accessed November 20, 2023, 2023. https://www.covid19treatmentguidelines.nih.gov/

25. Group RC, Horby P, Lim WS, et al. Dexamethasone in Hospitalized Patients with Covid-19. N Engl J Med. Feb 25 2021;384(8):693–704. doi:10.1056/NEJMoa2021436

26. Danesh V, White HD, Tecson KM, et al. Daily Oxygenation Support for Patients Hospitalized With SARS-CoV-2 in an Integrated Health System. Respir Care. Apr 2023;68(4):497–504. doi:10.4187/respcare.10401

27. Besnier E, Hobeika S S NS, et al. High-flow nasal cannula therapy: clinical practice in intensive care units. Ann Intensive Care. Sep 4 2019;9(1):98. doi:10.1186/s13613-019-0569-9

28. Li J, Albuainain FA, Tan W, Scott JB, Roca O, Mauri T. The effects of flow settings during high-flow nasal cannula support for adult subjects: a systematic review. Crit Care. Feb 28 2023;27(1):78. doi:10.1186/s13054-023-04361-5

29. Adjei S, Hong K, Molinari NM, et al. Mortality Risk Among Patients Hospitalized Primarily for COVID-19 During the Omicron and Delta Variant Pandemic Periods - United States, April 2020-June 2022. MMWR Morb Mortal Wkly Rep. Sep 16 2022;71(37):1182–1189. doi:10.15585/mmwr.mm7137a4

30. Kartsonaki C, Baillie JK, Barrio NG, et al. Characteristics and outcomes of an international cohort of 600 000 hospitalized patients with COVID-19. Int J Epidemiol. Apr 19 2023;52(2):355–376. doi:10.1093/ije/dyad012

