## Supplemental Data for "Quantifying Oxygen Demand by Patients Hospitalized with COVID-19 at a Large Safety-Net Hospital Using Multiple Methodologies"

**Table S1. Oxygen Flow (*Xn)* Formulae by Oxygen Delivery Device**

| **Oxygen Delivery Device** | ***Xn*** |
| --- | --- |
| Standard Flow (SFO) | *Xn = liters O_2_ per minute* |
| High Flow (HFO) | *Xn = (liters O_2_ per minute) x [(FiO_2_ - 0.21)/0.79]* |
| Positive Pressure Ventilation (PPV) | *Xn^*^ = [(minute ventilation + (bias flow x RR x expiratory time/60))] x [(FiO_2_ - 0.21)/0.79]* |
| To calculate oxygen demand volume over a time interval, *Xn* is multiplied by the time interval $(T_{\left( n+1 \right)}-T_{n})$. Total oxygen demand is calculated as the sum of oxygen demand volume for each time interval during a hospitalization. | |

^*^Bias flow is rated by manufacturer and differs by make and model of PPV device. For this analysis, bias flow was assumed to be 5 LPM.

RR=respiratory rate

**Table S2. Proportions of Patients Receiving Oxygen from Different Respiratory Support Devices by Severity Scenario for Calculator-Based Modeling**

| **Device** | **Scenario 1 (%)** | **Scenario 2 (%)** | **Scenario 3 (%)** | **Scenario 4 (%)** | **Scenario 5 (%)** |
| --- | --- | --- | --- | --- | --- |
| Nasal Cannula | 90 | 76 | 60 | 45 | 30 |
| Facemask | 1 | 3 | 5 | 6 | 9 |
| Facemask with reservoir | 1 | 3 | 5 | 7 | 9 |
| HFO | 4 | 9 | 15 | 21 | 26 |
| CPAP or NIPPV | 1 | 1 | 2 | 3 | 3 |
| Ventilator | 3 | 8 | 13 | 18 | 23 |
